# Investigating the genotoxicity of occupational pesticide exposures in Arab countries: Protocol of a systematic review and meta-analysis

**DOI:** 10.1101/2022.08.31.22279322

**Authors:** Moustafa Sherif, Khadija Ramadhan Makame, Linda Östlundh, Marilia Silva Paulo, Abderrahim Nemmar, Bassam R. Ali, Rami Al Rifai, Károly Nagy, Balázs Ádám

## Abstract

**Background:** Tens of millions of agricultural workers are directly exposed to pesticides through handling pesticide products, mostly in developing countries. Systematic data synthesis on the genotoxic consequences of such occupational exposures and their human health risks in agricultural settings is required in Arab countries. We aim to conduct a systematic review and, if possible, a meta-analysis to review published literature about the genotoxicity of occupational pesticide exposures in Arab countries, with the objectives of characterising the (1) prevalence rates of genotoxic pesticide exposures, (2) extent of genotoxic insults, (3) attributed risk factors, and (4) preventive measures against pesticide-induced genotoxic effects detectable by biomonitoring.

**Methods:** The research will follow the guidelines of the Preferred Reporting Items for Systematic Reviews and Meta-Analyses Protocols (PRISMA-P) statement. A comprehensive search will be conducted in November 2022 in the electronic databases PubMed (NLM), EMBASE (Elsevier), Web of Science (Clarivate), Scopus (Elsevier), and Agricola in addition to WHO Index Medicus for the Eastern Mediterranean (IMEMR). The search will be performed without any restrictions for publication years. A filter for English and Arabic language will be applied.

The systematic search will include agricultural workers over the age of 18 years, located in Arabic speaking countries of the Middle East and North Africa (MENA) region, occupationally exposed to pesticides inducing genotoxic insult detectable by biomonitoring.

Records identified in the search will be imported into the systematic review tool Covidence for blinded screening and selection by two reviewers independently. The reviewers will then extract data and conduct risk of bias assessment using the Navigation Guide RoB tool and the RoB-SPEO tool. The results will be synthetized narratively in summary tables, and, if findings allow, meta-analysis, including subgroup and sensitivity analysis, will be conducted on the prevalence of genotoxic pesticide exposures, and on the effect size of risk factors. The systematic review methodology does not require ethics approval.

**Discussion:** The systematic review will consider various types of pesticide exposures and genotoxicity biomarker assays to determine prevalence and extent of such occupational genotoxic insults, the correlation between genetic damage and various risk factors, e.g. work conditions, types of pesticides, environmental exposure routes, and the applied preventive measures. The review will provide gap-filling information about genotoxic pesticide exposures of agricultural workers in the local context, as well as it will contribute to our general knowledge on pesticide genotoxicity.

**Systematic review registration:** PROSPERO registration number: CRD42022314453.

## Introduction

Pesticides are types of agrochemicals extensively used around the globe to control “pests” including bacteria, fungi, weeds, snails, insects, rodents, and worms. Pesticides may be classified according to their target species into different groups, such as biocides, herbicides, nematicides, fungicides, rodenticides, acaricides, insecticides, molluscicides, repellents, and growth regulators (1). As reported in 2011, these pesticides belong to 1630 chemical substances but commercialized under 10,400 product names (2) and formulated in different forms, such as granules, liquids, concentrates, resin strips, impregnated pellet-tablets, dusts, and encapsulated particles (3). To increase pesticide efficacy and avoid pesticide resistance, these formulations are continuously developed and contain several further ingredients, such as surfactants and solvents that may constitute additional toxicity factors (4, 5).

Human exposure to pesticides can occur through various routes: oral intake of contaminated water or food, inhalation of aerosols, and skin absorption during mixing, loading, spraying, crop harvesting, and livestock management. At least 300 million people around the world are annually exposed to acute toxicity of pesticides in environmental and occupational settings (6), mainly in developing countries and especially agriculture workers who are involved in processing and application of these compounds in agricultural settings (3). Exposure to pesticides can be associated with high risk of developing many diseases depending mainly on the chemical properties of the pesticide, dose and exposure time. As previously reported, respiratory diseases, cancers, diabetes, immune toxicity and neurodegenerative and neurodevelopmental disorders may develop in the exposed populations (7). The toxicity to human and other non-target species may be due to their poor selectivity to target species and the similarities of all organisms in the basic biological processes. As a result, the International Agency for Research on Cancer (IARC) classified some pesticides, such as diazinon, glyphosate, and malathion (8, 9), being possible human carcinogens as shown in human epidemiological studies and experimental animal data, and the use of various pesticides were banned or strictly regulated (e.g. US-EPA, Dir. 91/414/EEC regulations).In Europe, the regulations are mainly based on the assessment of adverse effects of active ingredients along with a representative formulation. However, these regulations frequently fail to adequately estimate the potential long-term effects, including genotoxicity and carcinogenicity, and the interaction of the active and other ingredients of pesticide formulations.

Several agrochemical substances such as dichlorodiphenyltrichloroethane, commonly known as DDT were shown experimentally to be genotoxic and mutagenic, that may lead to the induction of cancer (10). For a better regulation and protection of exposed populations against risk for DNA damage-related chronic diseases and cancer, biomonitoring assessment tools have been developed to assess early effects of genotoxicity and mutagenicity. These biomarker assays are early disease indicators that can be used to investigate pesticide-exposed populations, as previously reviewed (7, 11, 12) and aim to measure various genetic endpoints, such as gene mutations, chromosomal alterations (micronuclei and aberrations), and direct DNA damage.

### Rationale

As far as we are aware, systematic review and synthesis of biological effect monitoring data on occupational pesticide exposure induced DNA damage among agricultural workers has not yet been done in Arab countries. Therefore, the goal of this systematic review is to retrieve and analyze publications from the Arab world on genotoxic biomonitoring studies in agriculture workers exposed to pesticides in occupational settings. The findings may allow for identifying gaps in knowledge and determine future directions in the development of preventive measures in Arab countries. The Arab World consists of 19 countries in the Middle East and North Africa: Algeria, Bahrain, Egypt, Iraq, Jordan, Kuwait, Lebanon, Libya, Morocco, Mauritania, Oman, Palestine, Qatar, Saudi Arabia, Sudan, Syria, Tunisia, the United Arab Emirates and Yemen.

### Objectives

The aim of the research is to conduct a systematic review by compiling and synthetizing the information presented in the published peer-reviewed literature regarding genotoxic pesticide exposures of agricultural workers in Arab countries, and to perform meta-analysis, if the results allow.

To accomplish this aim, the following set of objectives will be studied:

1. To systematically review of in extenso research papers published in scientific journals in English or Arabic on genotoxic occupational pesticide exposures among agriculture workers in Arab countries.
2. To review the prevalence of genotoxic occupational pesticide exposures among agriculture workers in Arab countries.
3. To review the extent of genetic damage potentially caused by occupational pesticide exposure in agricultural workers in Arab countries.
4. To determine statistically significant risk factors of genotoxic occupational pesticide exposure incidents.
5. To summarize the existing preventive measures used in Arab countries for controlling pesticide-induced genotoxic insult detectable by biomonitoring in agricultural settings.

Additionally, the following objectives will be accomplished if we can perform meta-analysis:

1. To calculate pooled frequency estimates for the prevalence of genotoxic pesticide exposures in Arab countries among agriculture workers, which could also allow for subgroup analysis.
2. To calculate pooled effect size estimates for potential risk factors of genotoxic occupational pesticide exposures, which could also allow for subgroup analysis.

## Materials and methods

### Protocol and registration

The protocol has been registered in the International Prospective Register of Systematic Reviews (PROSPERO) on 3rd March 2022 [ID: CRD42022314453]. This protocol follows the PRISMA-P statement that refers to the Preferred Reporting Items for Systematic Reviews and Meta-Analyses Protocols (13). The scheduled start date of the study is 1st November 2022 and will continue until 1st November 2024. The PRISMA-P checklist is accessible as a supplemental file (S1). The final review will go along with the new PRISMA 2020 statement (14, 15).

### Eligibility criteria

A population, exposure, comparator and outcomes (PECO) statement was established to review data on genotoxic occupational pesticide exposures occurring to agriculture workers in Arab countries.

#### 1. Participants/population

Inclusion: Adults (>18 years-old) who work as professional agriculture workers (farmers, including pesticide applicators), defined as people who are potentially exposed to pesticides during agricultural practices located in Arabic speaking countries of the Middle East and North Africa (MENA) region.

Exclusion: Population working in other sectors than agriculture, located outside the Arabic speaking countries of the MENA region, and less than 18 years old.

#### 2. Intervention(s), exposure(s)

Inclusion: Agriculture workers who are potentially exposed to pesticides during agricultural work activities.

Pesticide products include herbicides, insecticides, fungicides, biocides, defoliants, nematicides, acaricides, rodenticides, molluscicides, avicides, miticides and chemical repellents which are used in agricultural fields and animal farms to increase productivity and decrease losses.

Exclusion: Exposure to pesticides that is not occurring in agricultural settings, and exposure to any chemicals and genotoxic agents other than pesticides.

#### 3. Comparator/Control

There will be no comparator for the prevalence and extent of detectable DNA damage potentially attributable to occupational exposure to pesticides in agricultural workers in Arab countries.

The comparator for the identification and for the determination of the effect size of exposure to genotoxic pesticide groups, active ingredients, formulations and mixtures, as well as to contributing factors, will be populations not exposed directly to pesticides, e.g., workers employed in organic agricultural settings not applying pesticides or the general population.

#### 4. Outcome

Genetic damage measured by human biomonitoring using recognized genotoxicity tests such as DNA strand break measurements in cells (e.g., comet assay, alkaline elution, alkaline unwinding and hydroxyapatite chromatography), cytogenetic assays (micronucleus and chromosomal aberration assays, including the use of fluorescence in situ hybridization and chromosome painting), and mutagenicity assays.

Additional outcomes will be estimating the prevalence and identifiable risk factors of occupational genotoxicity among agriculture workers who are potentially exposed to pesticides in Arab countries, and evaluating the preventive measures used in practice to lower the risk of genotoxic occupational pesticide exposures in Arab countries.

### Types of studies

Observational studies (cross-sectional, case-control, and cohort studies) will be included in the review to determine the prevalence and identify risk factors of genotoxic pesticides exposure among agriculture workers in Arab countries. Only in extenso research articles publishing results of human biomonitoring will be considered. In vitro, in silico or animal studies, case reports, opinion articles, commentaries, letters, review articles, clinical trials, published abstracts and conference proceedings will not be used.

### Information sources

We will search systematically for studies that match our criteria and are published in peer-reviewed scientific journals in electronic databases including PubMed (NLM), EMBASE (Elsevier), Web of Science (Clarivate), Scopus (Elsevier), and Agricola in addition to WHO Index Medicus for the Eastern Mediterranean (IMEMR).

### Search strategy

We conducted preliminary searches in PubMed and Scopus in December 2021 – March 2022 based on the PECO statement with inclusion and exclusion criteria as defined above. Search terms identified with the help of the subject specialists were systematically developed with the help of PubMed’s MeSH and the search technical specifications advised and reviewed by an information specialist. The pre-search in PubMed and Scopus is available online in the supplemental file (S2).

A full systematic search of the literature in 6 databases (PubMed, EMBASE, Web of Science, Scopus, Agricola, Index Medicus for the Eastern Mediterranean) will be conducted in August 2022 based on the strategy developed in the pre-search. The search will be performed by applying all search terms in the databases MeSH/thesauruses and in the “title” and “abstract” fields. All databases will be searched from their inception. A filter for English and Arabic languages will be used. No additional filers or limitations will be applied for the best possible results and to capture all pre-indexed studies. The PRISMA S checklist for searching and reproducible search strings for all databases will be provided as an appendix in the final review.

The search will be updated in all databases ahead of completing the manuscript to ensure the inclusion of potential eligible studies published during the research process. Finally, the reference lists of the included studies will be screened manually by two independent reviewers.

### Study records and management

#### 1. Data management

Records located in the literature search will be uploaded to the systematic review software Covidence (Veritas Health Innovation) for title/abstract and full text screening, conflict resolving, selection, and data extraction. All modules in Covidence are blinded.

#### 2. Selection process

All records retrieved in the search will be automatically de-deduplicated and screened in Covidence (16). Two review authors will independently screen the titles and abstracts of the studies and assess them if they are potentially eligible according to the inclusion and exclusion criteria. The same reviewers together with the National Medical Library team at UAEU will then work to recover the full text of the selected studies. If there are any disagreement about inclusion of certain studies in either screening rounds, consensus will be determined by a third reviewer through the blinded conflict module in Covidence. PRISMA flow-diagram will be used to record the results of the screening and selection process, including reasons for full text exclusion (15). Finally, Cabell’s Predatory Reports will be used to confirm the academic quality of eligible studies published in open access journals.

### Data extraction

A data extraction template will be developed, and pilot tested on two articles in Excel, then populated in Covidence, where independent data extraction will be performed by two reviewers. In case of any discrepancies, a third reviewer will resolve it using the blinded conflict resolving function of the extraction module in Covidence. We might contact study authors to retrieve any missing data.

Extracted information will include publication data (title, DOI, year of publication, name of the first author, country), study settings (study type, period of data collection, risk factors and confounder adjustment, measurement of exposure and outcome, genotoxicity test(s) applied and measured endpoint(s) of genetic damage), study population (baseline characteristics), prevalence, average level and dispersion of measured genetic damage, and applied preventive measures. We will also collect information on conflict of interest and funding.

### Risk of bias (RoB) in individual studies

Two reviewers will individually assess the RoB in the eligible studies by the Navigation Guide RoB tool that is customized exclusively to occupational health systematic reviews (17). This tool determines RoB in domains of selection bias, ascertainment bias, accuracy of exposure and outcome evaluation, and selective reporting. Other biases and conflict of interest will be assessed with domains from the RoB-SPEO tool (Risk of Bias in Studies estimating Prevalence of Exposure to Occupational risk factors). This tool was developed by the World Health Organization (WHO) and the International Labour Organization (ILO) to be used in joint estimates of work-related burden of disease and injury (WHO/ILO Joint Estimates) (18). Any uncertainties over the risk of bias assessment between the two reviewers in particular studies will be decided by a third reviewer.

### Data synthesis

A narrative synthesis of the findings and summary tables will be provided from the selected eligible studies to summarize the types of pesticides, agriculture workers’ characteristics and types of outcomes. If findings allow, meta-analysis will be performed on the prevalence of genotoxic occupational pesticides exposures in Arab countries and on potential risk factors.

If two or more studies are found with appropriate estimates on outcome frequency and/or on risk factor effect, two reviewers will individually explore the clinical heterogeneity of the studies and combine them for meta-analysis.

If the combined studies are clinically sufficiently homogenous, the effect and/or frequency estimates will be pooled in a quantitative meta-analysis, using the inverse variance method with a random-effects model to account for cross-study heterogeneity (17). Statistical heterogeneity of the studies will be analyzed using the I2 statistics (19). The meta-analysis will be performed in RevMan software (20). The combined estimations will be shown in forest plots.

The funnel plot graphic representation will be used to visually evaluate the publication bias, and we will run a sensitivity analysis in case of the presence of outliers or asymmetry in the funnel plot.

### Analysis of subgroups or subsets

The discrepancies in frequency and/or effect estimates in the population will allow for subgroup analyses by the relevant variables or combination of variables (e.g. age, sex, country, job, agriculture sector or by a grouping of these variables). Sensitivity analyses will be conducted on studies judged as ‘low’ or ‘probably low’ in terms of RoB and conflict of interest.

### Quality of cumulative evidence

Quality of evidence will be assessed by all reviewers using the Grading of Recommendations Assessment, Development and Evaluation (GRADE) to assess individual risk for bias, inconsistency, indirectness, imprecision, and publication bias. GRADE scoring is based on the Navigation Guide quality of evidence assessment tool, which will allow us to provide summary-based evidence statements (21-23).

## Discussion

Pesticides used in agricultural and residential settings are deeply investigated experimentally and epidemiologically to evaluate their contribution to the development of degenerative and cancer diseases. However, the rapid development in the pesticide market in terms of chemical formulations has led to a high variability in the formulations, unreliable exposure data, and many confounding factors. The exposure to more than one class of pesticide resulted in poor assessment of cause-effect relationship that can be used to associate specific compounds with certain diseases.

Researchers globally used genotoxicity biomarkers as early indicators of risk for pesticide associated diseases in exposed human populations. We will perform a systematic review on the available data in the scientific literature that meet the aforementioned criteria. Previous reviews to analyze those studies were done but for other purposes (11, 12, 24). Despite the limitations and uncertainty, they all concluded that pesticides exposure is genotoxic.

As far as we are aware, systematic review and synthesis of biological effect monitoring data on occupational pesticide exposure-induced DNA damage among agricultural workers has not yet been carried out in Arab countries. As scientific debate is ongoing about the genotoxic and carcinogenic potential of several pesticide active ingredients and formulations, such a systematic review can provide a yet missing summarized evidence not only for the general scientific knowledge but also for the local occupational and environmental health and safety policymakers.

The reviewed information on pesticide exposures in agricultural workers and their effect on DNA damage will add valuable information to the scientific community and international professional bodies, and, for the first time, it informs regional and local policymakers about the exposure patterns and related genotoxicity in Arab countries.

The main limitations in this study could be the possible shortage of reliable publications and data on pesticide exposure and related genotoxic insult in Arab countries, the presence of confounding factors, especially when occupational workers are exposed to various types of pesticides and other genotoxic agents.

## Data Availability

All relevant data are within the manuscript and its Supporting Information files.

## Supporting information

**S1 File. PRISMA-P 2015 checklist**.

(DOCX)

**S2 File. Preliminary search**.

(DOCX)

### List of abbreviations

ILO: International Labour Organization.
MENA: Middle East and North Africa.
PRISMA: Preferred Reporting Items for Systematic Reviews and Meta-analyses.
PECO: population, exposure, comparator and outcomes.
RoB: Risk of bias.

## Declarations

### Ethics and Dissemination

Ethics approval is not necessary for such type of research (systematic review), because no public members or patients will participate in the study.

The review findings will be made publicly available in electronic format in a peer-reviewed scientific journal. Moreover, if the findings suggest a modification in pesticide management, a synopsis will be distributed in the East Mediterranean Region of the WHO to prominent agriculture and occupational policy makers.

## Author Contributions

MS, KRM, LÖ, MSP, AN, BA, RHA-R, KN, and BÁ were involved in conceptualization and protocol development. MS, LÖ and BÁ have developed the search strategy. Literature search will be conducted by MS and LÖ. Screening will be performed by MS and KRM, with BÁ resolving conflicts. MS and KRM will extract data, which will be validated by BÁ, KN, MSP, AN, BA, and RHA-R. RoB will be performed by MS and KRM, with BÁ resolving conflicts. Quality of evidence will be assessed by all reviewers.

## Patient consent for publication

Not applicable.

## Provenance and peer review

Not commissioned; externally peer reviewed.

## Supplemental material

This content has been supplied by the author(s). Any opinions or recommendations discussed are solely those of the author(s) and are not endorsed by PLOS ONE.

## Acknowledgements

We acknowledge the National Medical Library team at UAEU for providing technical support to develop the search strings for the study.

## Transparency statement

We are committed to operating responsibly and with high standards in this systematic review, and we will not omit any important aspects of the study that maybe relevant. We will clearly explain and discuss any discrepancies from this protocol in our publications.

## Amendments

Eventual minor changes to the protocol after publishing, will be transparently recorded with date and the rationale in the PROSPERO registration.

## Availability of data and materials

Since no datasets were created or analyzed at this stage, data sharing is not applicable. The data from this ongoing study can be found in this article or its supplemental materials. Once this study comes to an end, the study data will be released to the public.

## References

1. Food Safety: Pesticides European Commission: Directorate-General for Health and Food Safety [Available from: https://ec.europa.eu/food/plants/pesticides_en.

2. The Pesticide Manual. sixteenth ed. Hampshire, UK: British Crop Protection Council; 2011.

3. Bolognesi C, Holland N. The use of the lymphocyte cytokinesis-block micronucleus assay for monitoring pesticide-exposed populations. Mutation Research/Reviews in Mutation Research. 2016;770:183–203.

4. Clendennen SK, Boaz NW. Chapter 14 - Betaine Amphoteric Surfactants—Synthesis, Properties, and Applications. In: Hayes DG, Solaiman DKY, Ashby RD, editors. Biobased Surfactants (Second Edition): AOCS Press; 2019. p. 447–69.

5. Mustafa IF, Hussein MZ. Synthesis and Technology of Nanoemulsion-Based Pesticide Formulation. Nanomaterials (Basel). 2020;10(8):1608.

6. Jeyaratnam J. Acute pesticide poisoning: a major global health problem. World health statistics quarterly 1990; 43 (3): 139-144. 1990.

7. Bolognesi C. Genotoxicity of pesticides: a review of human biomonitoring studies. Mutat Res. 2003;543(3):251–72.

8. Bastos PL, Bastos A, Gurgel ADM, Gurgel IGD. Carcinogenicity and mutagenicity of malathion and its two analogues: a systematic review. Cien Saude Colet. 2020;25(8):3273–98.

9. Feulefack J, Khan A, Forastiere F, Sergi CM. Parental Pesticide Exposure and Childhood Brain Cancer: A Systematic Review and Meta-Analysis Confirming the IARC/WHO Monographs on Some Organophosphate Insecticides and Herbicides. Children (Basel). 2021;8(12).

10. WHO-JMPR Toxicological Monographs-Pesticide residues in food 2022 [Available from: https://inchem.org/pages/jmpr.html.

11. Bolognesi C, Creus A, Ostrosky-Wegman P, Marcos R. Micronuclei and pesticide exposure. Mutagenesis. 2011;26(1):19–26.

12. Bull S, Fletcher K, Boobis AR, Battershill JM. Evidence for genotoxicity of pesticides in pesticide applicators: A review. Mutagenesis. 2006;21(2):93–103.

13. Moher D, Shamseer L, Clarke M, Ghersi D, Liberati A, Petticrew M, et al. Preferred reporting items for systematic review and meta-analysis protocols (PRISMA-P) 2015 statement. Systematic Reviews. 2015;4(1):1.

14. edited by Julian PTH, Sally G. Cochrane handbook for systematic reviews of interventions: Chichester, West Sussex ; Hoboken NJ : John Wiley & Sons, [2008] ©2008; 2008.

15. Page MJ, McKenzie JE, Bossuyt PM, Boutron I, Hoffmann TC, Mulrow CD, et al. The PRISMA 2020 statement: an updated guideline for reporting systematic reviews. BMJ. 2021;372:71.

16. Covidence systematic review software. Melbourne, Australia: Veritas Health Innovation; 2021.

17. Woodruff TJ, Sutton P. The Navigation Guide systematic review methodology: a rigorous and transparent method for translating environmental health science into better health outcomes. Environ Health Perspect. 2014;122(10):1007–14.

18. Pega F, Norris SL, Backes C, Bero LA, Descatha A, Gagliardi D, et al. RoB-SPEO: A tool for assessing risk of bias in studies estimating the prevalence of exposure to occupational risk factors from the WHO/ILO Joint Estimates of the Work-related Burden of Disease and Injury. Environ Int. 2020;135:105039.

19. Higgins JP, Thompson SG, Deeks JJ, Altman DG. Measuring inconsistency in meta-analyses. Bmj. 2003;327(7414):557–60.

20. Review manager (revman) software. 5.3 ed. Copenhagen, Denmark: The Nordic Cochrane Centre: The Cochrane Collaboration; 2014.

21. Schünemann HJ, Oxman AD, Vist GE, Higgins JP, Deeks JJ, Glasziou P, et al. Chapter 12: Interpreting results and drawing conclusions. Cochrane handbook for systematic reviews of interventions: The Cochrane Collaboration; 2011.

22. Morgan RL, Thayer KA, Bero L, Bruce N, Falck-Ytter Y, Ghersi D, et al. GRADE: Assessing the quality of evidence in environmental and occupational health. Environ Int. 2016;92-93:611–6.

23. Lam J, Sutton P, Padula AM, Cabana MD, Woodruf TJ. Applying the navigation guide systematic review methodology case study #6: association between formaldehyde exposure and asthma: a systematic Review of the evidence: protocol. San Francisco, CA: University of California at San Francisco. 2016.

24. Yang HY, Feng R, Liu J, Wang HY, Wang YD. Increased frequency of micronuclei in binucleated lymphocytes among occupationally pesticide-exposed populations: a meta-analysis. Asian Pac J Cancer Prev. 2014;15(16):6955–60.

