## Supplemental file S2 for "Investigating the genotoxicity of occupational pesticide exposures in Arab countries: Protocol of a systematic review and meta-analysis"

### Preliminary literature search in PubMed and Scopus

| Source and search date | Search string | Results and Notes |
|---|---|---|
| **PubMed** (NLM)<br><br>**Coverage:** from database inception to search date<br><br>**Search date:** 2022-03-29 | (((agrochemical*[Title/Abstract] OR "chemical*"[Title/Abstract] OR agrichemical*[Title/Abstract] OR "plant protection product*"[Title/Abstract] OR pesticid*[Title/Abstract] OR biocid*[Title/Abstract] OR herbicid*[Title/Abstract] OR weedkiller*[Title/Abstract] OR "weed killer*"[Title/Abstract] OR defolian*[Title/Abstract] OR insecticid*[Title/Abstract] OR nematicid*[Title/Abstract] OR molluscicid*[Title/Abstract] OR piscicid*[Title/Abstract] OR avicid*[Title/Abstract] OR rodenticid*[Title/Abstract] OR bactericid*[Title/Abstract] OR repellent*[Title/Abstract] OR antimicrob*[Title/Abstract] OR "antiparasit*"[Title/Abstract] OR fungicid*[Title/Abstract] OR lampricid*[Title/Abstract] OR acaricid*[Title/Abstract] OR miticid*[Title/Abstract] OR "mite control*"[Title/Abstract] OR algicid*[Title/Abstract] OR algaecid*[Title/Abstract] OR chemosterilant*[Title/Abstract] OR "Agrochemicals"[MeSH:NoExp] OR "Pesticides"[MeSH] OR "Antiparasitic Agents"[MeSH] OR "Insect Repellents"[MeSH]) AND ("Poisons"[Mesh:NoExp] OR poison*[Title/Abstract] OR toxic*[Title/Abstract] OR toxicogenetic*[Title/Abstract] OR "Toxicogenetics"[MeSH] OR genotox*[Title/Abstract] OR cytotox*[Title/Abstract] OR "Cytotoxins"[MeSH] OR antimetabolite*[Title/Abstract] OR "Antimetabolites"[MeSH] OR antispermat*[Title/Abstract] OR "Antispermatogenic Agents"[MeSH] OR cardiotox*[Title/Abstract] OR "Cardiotoxins"[MeSH] OR dermatotox*[Title/Abstract] OR dermotox*[Title/Abstract] OR "Dermotoxins"[MeSH] OR hepatotox*[Title/Abstract] OR nephrotox*[Title/Abstract] OR pneumotox*[Title/Abstract] OR immunotox*[Title/Abstract] OR "Immunotoxins"[MeSH] OR neurotox*[Title/Abstract] OR "Neurotoxins"[MeSH] OR "Toxic Actions"[MeSH] OR noxae*[Title/Abstract] OR hazard*[Title/Abstract] OR "Toxicity Tests"[MeSH] OR "Mutagenicity Tests"[MeSH] OR "Carcinogenicity Tests"[MeSH] OR pharmacogenomic*[Title/Abstract] OR "Pharmacogenomic Testing"[MeSH] OR "Comet Assay"[MeSH] OR "comet assay*"[Title/Abstract] OR "Micronucleus Tests"[MeSH] OR "micronucleus test*"[Title/Abstract] OR "chromosome aberrat*"[Title/Abstract] OR "chromosomal aberrat*"[Title/Abstract] OR "Chromosome Aberrations"[MeSH] OR denaturation*[Title/Abstract] OR "Nucleic Acid Denaturation"[MeSH] OR "fluorescent in situ hybridization*"[Title/Abstract] OR "In Situ Hybridization, Fluorescence"[MeSH] OR "Nucleic Acid Hybridization"[MeSH] OR "nucleic acid hybridization*"[Title/Abstract] OR "ames test*"[Title/Abstract] OR aneuploidy*[Title/Abstract] OR "Aneuploidy"[Mesh] OR topoisomerase*[Title/Abstract] OR "DNA | **Results:** 1,354<br><br>**Notes:**<br>All search terms are searched in the fields: “title” and “abstract” and in the MeSH (when available).<br><br>Filters for English and Arabic languages are applied. |

| | |
|---|---|
| | Topoisomerases"[Mesh] OR "Teniposide"[Mesh] OR teniposide*[Title/Abstract] OR "Etoposide"[Mesh] OR etoposide*[Title/Abstract] OR greenscreen*[Title/Abstract] OR "γH2AX*"[Title/Abstract] OR "pH2AX*"[Title/Abstract] OR "high content screening*"[Title/Abstract] OR "pH3"[Title/Abstract] OR "cycle arrest*"[Title/Abstract] OR phospho-histon*[Title/Abstract] OR phosphohiston*[Title/Abstract] OR caspase*[Title/Abstract] OR "Caspases"[MeSH] OR "tubulin microtubule*"[Title/Abstract] OR "profiling assay*"[Title/Abstract] OR steatosis*[Title/Abstract] OR "Genetic Carrier Screening"[MeSH] OR carcinogen*[Title/Abstract] OR "Carcinogens"[MeSH] OR "Carcinogenesis"[MeSH] OR mutagen*[Title/Abstract] OR "Mutagens"[MeSH] OR "Mutagenesis"[MeSH] OR mutation*[Title/Abstract] OR "Mutation"[MeSH] OR teratogen*[Title/Abstract] OR "Teratogens"[MeSH] OR "Teratogenesis"[MeSH] OR disorder*[Title/Abstract] OR genetic*[Title/Abstract] OR "DNA"[Title/Abstract] OR "DNA"[Mesh] OR "RNA"[Title/Abstract] OR "RNA"[Mesh] OR "DNA Damage"[MeSH] OR damag*[Title/Abstract] OR insult*[Title/Abstract] OR adduct*[Title/Abstract] OR alkylation*[Title/Abstract] OR "Alkylation"[MeSH] OR alkylating[Title/Abstract] OR methylation*[Title/Abstract] OR "Methylation"[MeSH] OR oxidizing[Title/Abstract] OR oxidant*[Title/Abstract] OR "Oxidative Stress"[MeSH] OR "oxidative stress*"[Title/Abstract] OR "free radical*"[Title/Abstract] OR "Free Radicals"[MeSH] OR "Cell Survival"[MeSH] OR viabilit*[Title/Abstract] OR viable*[Title/Abstract] OR necro*[Title/Abstract] OR "Necrosis"[MeSH] OR apopto*[Title/Abstract] OR "Apoptosis"[MeSH] OR "Genes, Lethal"[MeSH] OR lethal*[Title/Abstract] OR "Tissue Survival"[MeSH] OR "surviv*"[Title/Abstract] OR "adverse effect*"[Title/Abstract] OR "side effect*"[Title/Abstract] OR "Long Term Adverse Effects"[MeSH] OR harmful[Title/Abstract] OR disease*[Title/Abstract] OR "Disease"[MeSH] OR illness*[Title/Abstract] OR syndrome*[Title/Abstract] OR "Syndrome"[MeSH] OR symptom*[Title/Abstract] OR abnormal*[Title/Abstract] OR irritant*[Title/Abstract] OR "Irritants"[MeSH] OR cancer*[Title/Abstract] OR neoplas*[Title/Abstract] OR "Neoplasms"[MeSH] OR tumor*[Title/Abstract] OR tumour*[Title/Abstract] OR malignan*[Title/Abstract] OR carcinoma*[Title/Abstract] OR "Carcinoma"[MeSH] OR malformat*[Title/Abstract] OR anomal*[Title/Abstract] OR abnormal*[Title/Abstract] OR "congenital defect*"[Title/Abstract] OR "birth defect*"[Title/Abstract] OR "Congenital Abnormalities"[MeSH] OR reprotox*[Title/Abstract] OR reproduct*[Title/Abstract] OR "Reproduction"[MeSH] OR allerg*[Title/Abstract] OR "Allergy and Immunology"[MeSH] OR hypersensitiv*[Title/Abstract] OR "Hypersensitivity"[MeSH] OR histolog*[Title/Abstract] OR "Histology"[MeSH] OR endocrin*[Title/Abstract] OR "Endocrine System"[MeSH] OR |

| | |
|---|---|
| | "Endocrine Disruptors"[MeSH] OR neuroendocrin*[Title/Abstract] OR "neuro-endocrin*"[Title/Abstract] OR "Neurosecretory Systems"[MeSH] OR physiopath*[Title/Abstract] OR pathophysi*[Title/Abstract] OR patho*[Title/Abstract] OR "Pathology"[MeSH]) AND ("Bahrain"[MeSH] OR Bahrain*[Title/Abstract] OR "Iraq"[MeSH] OR Iraq*[Title/Abstract] OR "Jordan"[MeSH] OR Jordan*[Title/Abstract] OR "Kuwait"[MeSH] OR Kuwait*[Title/Abstract] OR "Lebanon"[MeSH] OR Lebanon[Title/Abstract] OR Lebanese*[Title/Abstract] OR "Oman"[MeSH] OR Oman*[Title/Abstract] OR "Qatar"[MeSH] OR Qatar*[Title/Abstract] OR "Saudi Arabia"[MeSH] OR Saudi*[Title/Abstract] OR KSA[Title/Abstract] OR "Syria"[MeSH] OR Syria*[Title/Abstract] OR "United Arab Emirates"[MeSH] OR "United Arab Emirates"[Title/Abstract] OR UAE[Title/Abstract] OR Emirat*[Title/Abstract] OR "Yemen"[MeSH] OR Yemen*[Title/Abstract] OR "Egypt"[MeSH] OR Egypt*[Title/Abstract] OR "Sudan"[MeSH] OR "Sudan*"[ Title/Abstract] OR "Mauritania"[MeSH] OR "Mauritan*"[Title/Abstract] OR Maghreb*[Title/Abstract] OR Maghrib*[Title/Abstract] OR Morocco*[Title/Abstract] OR Morocco [MeSH] OR Algeri*[Title/Abstract] OR Algeria[MeSH] OR Libya*[Title/Abstract] OR Libya[MeSH] OR Tunis*[Title/Abstract] OR Tunisia[MeSH] OR Palestin*[Title/Abstract] OR "Arab World"[MeSH] OR "Africa, Northern"[MeSH] OR "Middle East*"[Title/Abstract] OR "Middle East"[MeSH:NoExp] OR "Near East"[Title/Abstract] OR "East Mediterranean"[Title/Abstract] OR "Eastern Mediterranean"[Title/Abstract] OR Arabic[Title/Abstract] OR Arabs[Title/Abstract] OR Arab[Title/Abstract] OR MENA[Title/Abstract] OR "Arabian Peninsula"[Title/Abstract] OR "North Africa*"[Title/Abstract] OR "Northern Africa*"[Title/Abstract] OR "West Bank*"[Title/Abstract] OR "Gaza Strip"[Title/Abstract] OR Levant*[Title/Abstract] OR gulf*[Title/Abstract] OR "Arabs"[MeSH] OR "Arab World"[MeSH]) AND (farmer*[Title/Abstract] OR "farm"[Title/Abstract] OR farms[Title/Abstract] OR ranch[Title/Abstract] OR rancher*[Title/Abstract] OR agronomist*[Title/Abstract] OR smallholder*[Title/Abstract] OR grazier*[Title/Abstract] OR farmhand*[Title/Abstract] OR "Farmers"[MeSH] OR agricultur*[Title/Abstract] OR grower*[Title/Abstract] OR reaper*[Title/Abstract] OR breeder*[Title/Abstract] OR cropper*[Title/Abstract] OR cultivator*[Title/Abstract] OR feeder*[Title/Abstract] OR gardener*[Title/Abstract] OR gleaner*[Title/Abstract] OR harvester*[Title/Abstract] OR horticulturist*[Title/Abstract] OR planter*[Title/Abstract] OR tiller*[Title/Abstract] OR rural*[Title/Abstract] OR "Rural Population"[MeSH] OR "worker*"[Title/Abstract] OR "labor*"[Title/Abstract] OR "labour*"[Title/Abstract]))) |

| | | |
|---|---|---|
| **Scopus** (Elsevier)<br><br>**Coverage:** from database inception to search date<br><br>**Search date:** 2022-03-29 | (TITLE-ABS-KEY ( farmer* OR "farm" OR farms OR ranch OR rancher* OR agronomist* OR smallholder* OR grazier* OR farmhand* OR "farmers" OR agricultur* OR grower* OR reaper* OR breeder* OR cropper* OR cultivator* OR feeder* OR gardener* OR gleaner* OR harvester* OR horticulturist* OR planter* OR tiller* OR rural* OR "worker*" OR "labor*" OR "labour*" ) ) AND ( TITLE-ABS-KEY ( "iraq*" OR "lebanon" OR "sudan" OR "mauritania" OR "middle east*" OR "near east" OR "east mediterranean" OR "eastern mediterran*" OR arabic OR arabs OR arab OR mena OR "arabian peninsula" OR "north africa*" OR "northern africa*" OR maghreb* OR maghrib* OR morocco* OR egypt* OR jordan* OR lebanes* OR syria* OR algeri* OR libya* OR tunis* OR uae OR "united arab*" OR emirat* OR saudi* OR ksa OR qatar* OR oman* OR yemen* OR kuwait* OR bahrain* OR palestin* OR "west bank" OR "gaza strip" OR levant* OR gulf* ) ) AND ( TITLE-ABS-KEY ( poison* OR toxic* OR genotox* OR cytotox* OR antimetabolite* OR antispermatogen* OR cardiotox* OR dermatotox* OR dermotox* OR reprotox* OR hepatotox* OR nephrotox* OR pneumotox* OR immunotox* OR neurotox* OR noxae* OR hazard* OR pharmacogenomic* OR "comet assay*" OR "micronucleus*" OR "aberrat*" OR "chromosom*" OR denaturation* OR "hybridization*" OR "ames" OR aneuploid* OR topoisomeras* OR teniposid* OR etoposid* OR greenscreen* OR "γh2ax*" OR "ph2ax*" OR "high content screening*" OR "ph3" OR "cycle arrest*" OR phospho-histon* OR phosphohiston* OR caspase* OR "tubulin microtubule*" OR "profiling assay*" OR steatosis* OR carcino* OR mutagen* OR "mutation" OR teratogen* OR disorder* OR genetic* OR "dna" OR "rna" OR damag* OR insult* OR adduct* OR alkylat* OR "methylation" OR oxidizing OR oxidant* OR "oxidative stress*" OR "free radical*" OR viabilit* OR viable* OR necro* OR "necrosis" OR apopto* OR lethal* OR "surviv*" OR "adverse effect*" OR "side effect*" OR harmful* OR disease* OR illness* OR syndrome* OR symptom* OR abnormal* OR irritant* OR cancer* OR neoplas* OR tumor* OR tumour* OR malignan* OR malformat* OR anomal* OR abnormal* OR "congenital*" OR "birth defect*" OR reproduct* OR allerg* OR hypersensitiv* OR histolog* OR endocrin* OR neuroend* OR "neuro-endocrin*" OR "neurosecret*" OR physiopatho* OR pathophysi* OR patholog* ) ) AND ( TITLE-ABS-KEY ( agrochemical* OR "chemical*" OR "plant protect*" OR pesticide* OR biocid* OR herbicid* OR weedkiller* OR "weed killer*" OR defoliant* OR insecticid* OR nematicid* OR molluscicid* OR piscicid* OR avicid* OR rodenticid* OR bactericid* OR repellent* OR antimicrob* OR "antiparasit*" OR | **Results:** 3,260<br><br>**Notes:**<br>All search terms are searched in the fields: “title”, “abstract” and “keywords” (here marked with “TITLE-ABS-KEY”)<br><br>No thesaurus available.<br><br>Filters for English and Arabic languages are applied. |

| | |
|---|---|
| | fungicid* OR lampricid* OR acaricid* OR miticid* OR "mite control*" OR algicid* OR algaecid* OR chemosterilant* ) ) |
